# Distinct serum anti-Aβ antibody patterns in hemorrhagic and inflammatory cerebral amyloid angiopathy manifestations

**DOI:** 10.1101/2020.10.07.20208330

**Authors:** Yannick Chantran, Jean Capron, Diana Doukhi, Johanna Felix, Mélanie Féroul, Florian Kruse, Thomas Chaigneau, Guillaume Dorothée, Thibault Allou, Xavier Ayrignac, Zina Barrou, Thomas de Broucker, Corina Cret, Guillaume Turc, Roxane Peres, Anne Wacongne, Marie Sarazin, Dimitri Renard, Charlotte Cordonnier, Sonia Alamowitch, Pierre Aucouturier

## Abstract

**Objective:** To study blood anti-Aβ antibodies in the context of spontaneous inflammatory or hemorrhagic CAA manifestations, which are similar to complications occurring after monoclonal anti-Aβ antibody immunotherapies.

**Methods:** In this case-control study, serum anti-Aβ antibody isotype, concentration, avidity, and reactivity toward soluble or fibrillary Aβ_1-40_ and Aβ_1-42_ isoforms were assessed using an ELISA-based multiplex analysis. Anti-Aβ serologic patterns were defined in CAA and CAA subgroups using multivariable logistic regression analyses.

**Results:** Fourty-one healthy aged controls and 64 CAA patients were recruited: 46 with hemorrhagic features (CAA-he) and 18 with CAA-related inflammation (CAA-ri). As compared to controls, the most striking features of CAA-related serological profiles were the following: i) both CAA-he and CAA-ri patients displayed lower binding diversity of anti-soluble Aβ_1-40_ IgM; ii) CAA-he patients displayed higher anti-soluble Aβ_1-40_ / fibrillary Aβ_1-42_ IgG4 concentrations ratio and higher anti-soluble Aβ_1-42_ IgG4 and IgA avidity; iii) CAA-ri patients displayed higher binding diversity of anti-soluble Aβ_1-40_ IgG3 and higher anti-fibrillary/soluble Aβ_1-42_ IgG4 dilution curve steepness ratio.

**Conclusion:** This proof-of-concept study revealed anti-Aβ antibody variations in CAA patients, some of which were associated to CAA clinical phenotypes, unveiling pathophysiological insights regarding CAA-hemorrhagic and inflammatory related events.

## Introduction

Cerebral amyloid angiopathy (CAA) relates to cortical and leptomeningeal vessel microangiopathy with accumulation of vascular amyloid fibrils made of amyloid-β peptide (Aβ).^1^ CAA is frequent in both Alzheimer’s disease (AD) and non-AD aged participants. Intracerebral hemorrhage (ICH) related to CAA (CAA-he) is a major health concern.^2^ CAA-related inflammation (CAA-ri) is a rare but treatable Aβ-related CNS vasculitis.^3^

Dose-dependent adverse events similar to CAA-he and CAA-ri were observed upon anti-Aβ immunotherapy in AD,^4^ suggesting a role of anti-Aβ antibodies in CAA manifestations. Subsequent investigations revealed elevated CSF levels of anti-Aβ IgG during CAA-ri.^5,6^ However, it was shown experimentally that blood-borne more than CSF-borne anti-Aβ antibodies can aggravate CAA manifestations in mouse models.^7,8^

As well as other auto-antibodies involved in neurodegenerative disorders, including anti-Tau protein^9^ and anti-alpha synuclein antibodies,^10^ anti-Aβ antibodies belonging to the “natural” auto-antibody repertoire: they are present in both diseased and healthy individuals, displaying low-avidity and multi-reactivity, and circulating mostly as immune complexes.^11^ These complexes require dissociation prior to analysis, in order to reveal serological specificities in AD.^11,12^ These features have likely hindered consistent analyses regarding anti-Aβ antibodies, which was also the case for anti-alpha synuclein antibodies.^10^ In AD, anti-Aβ antibodies measurement led to inconsistent conclusions.^11-14^ Circulating anti-Aβ antibodies were scarcely studied in CAA.

Our working hypothesis was that CAA and related clinical manifestations would relate to particular characteristics of serum anti-Aβ antibodies, including concentration, avidity, specific reactivity towards Aβ isoforms, and class and subclass diversity.

## Materials and methods

### Study design and participants

This case-control study enrolled participants from Jan. 2013 to Jun. 2019, through nine French medical centers: CAA patients, and healthy aged controls lacking clinical and MRI features of CAA. Patients with CAA were classified into: CAA-he inpatients with probable or definite CAA according to the modified Boston criteria;^15,16^ CAA-ri inpatients fulfilling the criteria for non-invasive diagnosis of CAA-ri.^17^ Demographics, medical history, and cerebral MRI were recorded at admission.

For the control group, inclusion criteria were: age > 55 years; recent MRI with normal diffusion sequences (transient ischemic stroke, functional neurological symptoms, or symptoms of peripheral origin); exclusion criteria were: cognitive decline (short Informant Questionnaire on Cognitive Decline in the Elderly (IQCODE-R) > 3.4); MRI-proven lobar ICH, recent ischemic stroke, or Fazekas grading ≥ 2 for white matter hyperintensities.

All eligible patients who gave their consent to participate in the study were included. All serum aliquots were kept at −20°C until use, with a median storage duration of 19 (3–42), 16 (3–50), and 27 (4–55) months (range) for the control, CAA-he, and CAA-ri groups, respectively.

### Standard Protocol Approvals, Registrations, and Patient Consents

The study protocol was approved by the ethics committee “Paris Ile de France V”. All participants representative were provided oral and written information, and gave oral or written consent to participate.

### Imaging assessment

Diagnoses were adjudicated by one board-certified neurologist (JC) specialized in stroke, blind to biological information. Cerebral lobar microhemorrhages were defined as small round foci of hypo-intense signal in T2*-GRE-weighted images, 10 mm or less in brain parenchyma and rated according to the microhemorrhages anatomical rating scale.^18^ Cortical superficial siderosis (cSS) was assessed according to the cSS multifocality scale.^19^

### Aβ preparations

Synthetic (>95%) Aβ_1–40_ and Aβ_1–42_ peptides (Proteogenix, Schiltigheim, France) were dissolved in hexafluoroisopropanol, and 450 μg aliquots were evaporated in low retention tubes and stored at −20 °C until use. Before use, lyophilized Aβ was dissolved in 10μL dimethylsulfoxide (DMSO), sonicated for 3 min at 300 Watts. For soluble preparations, aliquots were then mixed with 90 μL 30mM HEPES 10eq Cu^2+^ pH 7.4 buffer with 10mM or 160mM NaCl for Aβ_1-42_ and Aβ_1-40_, respectively. For fibril preparations, aliquots were then mixed with 90 μL 0.01N HCl or Aβ_1-40_ coating buffer, incubated at 37°C during 72h or 15 days, for Aβ_1-42_ and Aβ_1-40_, respectively.

### Multiplex ELISA for anti-Aβ antibody analyses

Freshly prepared soluble or fibrillar Aβ_1-40_ and Aβ_1-42_ (hereafter termed s-Aβ_40_, s-Aβ_42_, f-Aβ_40_ or f-Aβ_42_) were diluted to 15 μg/mL in coating buffer (30mM HEPES 160mM or 10mM NaCl (for Aβ_1–40_ and Aβ_1–42_, respectively) 10eq Cu^2+^ (for monomers) pH 7.4), distributed at 100 μL per well into ELISA plates (Greiner BioOne) and incubated 16 hours at 4°C. Serial dilutions of serum samples at 1:50 to 1:12800 in 0.1M Glycine-HCl buffer pH 3.0, were left 40 minutes at 20°C for dissociation of immune complexes, neutralized to pH 7.4 by adding the same volume of 2xPBS 4% BSA 0.02N NaOH, then 100μL were immediately deposited into s-Aβ_40_, s-Aβ_42_, f-Aβ_40_ or f-Aβ_42_-coated ELISA plates and incubated 1h at 20°C. After eight washes with PBS 0.05% Tween-20, bound antibodies of each IgG subclass were detected by 16h incubation at 4°C of monoclonal anti-human IgG1, IgG2, IgG3, or IgG4 antibodies (clones NL16, GOM2, ZG4 and RJ4, respectively). After eight washes with PBS 0.05% Tween-20, antibodies belonging to IgG, IgA and IgM classes and IgG subclasses were revealed after 1h incubation at 20°C with peroxidase-conjugated antisera (anti-mouse IgG for IgG subclasses, or anti-human IgG, IgA, and IgM, 1:5000 in washing buffer, Jackson ImmunoResearch Inc.). Washed plates were revealed with H_2_O_2_/o-phenylene-diamine substrate in 0.15 M urea buffer pH 5.0, the reaction stopped with 2N H_2_SO_4_, and optical densities (OD) measured at 492 nm. Non-specific signals obtained in uncoated wells were subtracted from overall signals retain OD relating to specific anti-Aβ binding. Of note, IgG2 antibodies yielded low binding to Aβ and unusable dilution curves as previously found in 33 AD and controls.^12^

### Quality management of multiplex ELISA

Samples were randomized and analyzed blindly to minimize bias due to manipulator and inter-assay variability. Randomization was stratified in order to include samples from all clinical groups, so any experimental bias would affect all groups comparably. Pools of human sera were used as internal standard for inter-experiment normalization, and internal control to assess inter-assay variability. Results over 2.5 standard deviation (sd) for one curve parameter were considered invalid.

### Determination of dilution curve parameters

Serum serial dilution curves follow a sigmoid-shaped signal in semi-logarithmic units, as illustrated on Fig 1A.^20^ The best fitting sigmoid curve parameters were determined by non-linear least square approach, following the equation:

**Figure 1.**
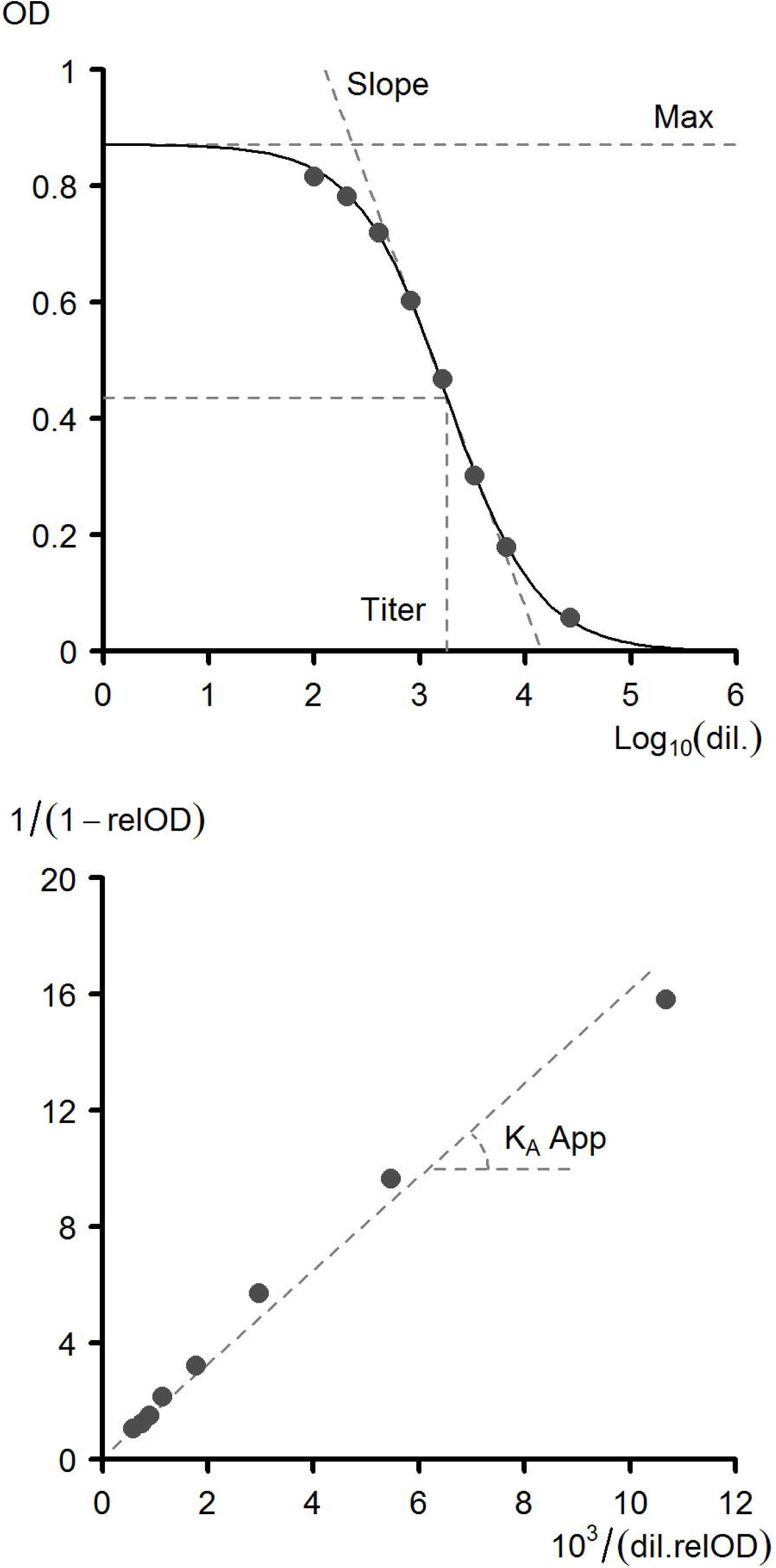
Determination of dilution curve parameters by sigmoid modeling and linearization procedure. A. Dilution curve obtained from a human serum sample following acidic dissociation of circulating immune complexes and neutralization, incubated on coated soluble Aβ1-42 and revealed with anti-IgG secondary antibody. The dashed thick line represents the sigmoid modeling of the curve, accurately described by i) the y-axis value of the left-sided plateau (*Maximum*); ii) the x-axis value at the inflexion point (*Titer*); iii) the slope at the inflexion point (*Steepness*). B. Linearization of the same experimental points and sigmoid model, which allows the determination of the apparent constant of avidity.

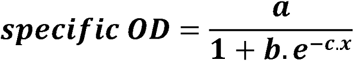

The values of *a, b*, and *c* directly relate to sigmoid curve parameters, hence to antibody properties. *Maximum* (*a*) signal obtained in antibody excess relate to the number of antigen binding sites, hence on the diversity of epitope recognition by polyclonal anti-Aβ antibodies. *Titer* (*ln(b)/c*) reflects the x-axis position of the curve, and depends on both concentration and avidity of the antibodies. The *Steepness* or *Slope* of the curve at the inflexion point (*–c/4a)* can vary with cooperativity phenomena occurring between distinct antibody binding sites. The apparent *Avidity* constant was calculated through a linearization procedure of the sigmoid curve (Fig 1B).^21^ In order to ensure appropriate goodness-of-fit of experimental data, only curves with R > 0.9 were taken into account.

### Statistical analysis

For each individual, 96 serological variables were analyzed (*Maximum, Titer, Steepness* and *Avidity* for anti-soluble and fibrillar Aβ40 and Aβ42 IgG, IgA, IgM, IgG1, IgG3, and IgG4). In order to limit multiple comparison bias, we chose not to perform univariate analysis. Multiple logistic regression models were computed using stepwise variable selection. The criteria for retaining a variable into the model were that all variables were significantly associated with the risk of belonging to the diseased group (tested by the Wald’s test; p<0.05) and that introducing this variable allowed a significant improvement of the model against the (k-1) model, as measured by a significant drop of the residual variance (tested by a Likelihood Ratio test; p-value < 0.05). Variance Inflation Factors (VIFs) were computed for each final model to ensure the absence of collinearity between variables. A VIF value of 1 is obtained when there is no collinearity between variables, while a VIF value > 5 witnesses high multicollinearity in the model. Wilcoxon’s test was used to compare the predicted response of the models between clinical groups. All statistical analyses were done with R version 3.6.1. The MASS package was used for model selection, and the beeswarm package for plots.

### Data Availability Statement

Data are available upon reasonable request.

## Results

### Patient demographics and clinico-radiological data

The study enrolled 105 participants: 41 healthy aged controls, 46 CAA-he patients, and 18 CAA-ri patients. Of note, eight patients from the CAA-he group had cSS but no ICH; their symptoms were : isolated cognitive decline in 3, transient focal episodes in 4 (2 with acute subarachnoidal hemorrhage); asymptomatic in one. Median (range) ages were 72 (55–89), 79 (59–90), 75 (64–87) years old in the control, CAA-he and CAA-ri groups, respectively. Male/Female ratios were 22/18, 24/21, and 8/9 in the control, CAA-he and CAA-ri groups, respectively. The main clinical and imaging findings in CAA-he and CAA-ri patients are presented in Table I.

### Quality assessment of the multiplex ELISA

The sigmoid modeling of experimental dilution curves showed excellent overall goodness-of-fit (mean R^2^ = 0.97; range stratified by antigen isoform: [0.97–0.98]; range stratified by antibody isotype: [0.95–0.99]). This confirmed that experimental dilution curves are appropriately described by the sigmoid model. Internal control mean coefficients of variation (CV) were inferior to 20% for all four parameters of all antibody isotypes and all Aβ isoforms (mean CV: 16%, 7%, 16%, and 12%, for *maxima, titer, steepness*, and *avidity*, respectively). This validated the multiplex ELISA as a reliable and reproducible method for assessing anti-Aβ antibody features.

### Serum anti-Aβ antibody patterns associated with CAA

Table II presents the anti-Aβ serologic parameters associated with CAA against controls. These parameters are those included in the multivariable logistic regression model after stepwise variable selection. Fig 2A presents the predicted response resulting from this CAA-model applied to each individual. This predicted response is associated with the probability of presenting CAA or not, and is modeled using the individual anti-Aβ profile regarding parameters presented in Table 2. In this complex profile, a higher diversity and concentration of anti-s-Aβ_40_ IgG3, a higher avidity of anti-s-Aβ_40_ IgG4 and a higher steepness of anti-f-Aβ_42_ IgG4 were associated with an increased probability of belonging to the CAA group. A lower diversity of anti-s-Aβ_40_ IgM, a lower avidity of anti-s-Aβ_42_ IgA, and a lower concentration of anti-f-Aβ_42_ IgG1 were also associated with an increased probability of belonging to the CAA group. All these variables, contributed independently to the model, without multicollinearity (all Variance Inflation Factors (VIFs) < 1.8).

**Table I.** Clinical and radiological characteristics of patients with cerebral amyloid angiopathy

|  |  |
| --- | --- |
| **CAA-ri patients (n=18)** |  |
| *Onset < 1 month* | 8 |
| *Symptoms* |  |
| Cognitive decline | 17 |
| Neurologic deficit | 11 |
| Epilepsy | 6 |
| Headache | 5 |
| *Histological diagnosis* | 2 |
| *MRI findings* |  |
| Asymetric white matter hypersignal reaching U fibers | 17 |
| Meningeal gadolinium enhancement (n=14 injected MRIs) | 6 |
| *Treatment* |  |
| Steroid alone | 11 |
| Steroid + Cyclophosphamid | 6 |
| None<sup>a</sup> | 3 |
| *Outcome (n=17)<sup>b</sup>* |  |
| Clinical and radiological improvement | 13 |
| Radiological improvement alone | 3 |
| **CAA-he patients (n=46)** |  |
| *MRI findings* |  |
| lobar micro-hemorrhages | 45 |
| cSS score (0-1-2-3-4) | 13/15/6/7/5 |
| Fazekas score (1-2-3) | 9/21/16 |
| *Modified Boston criteria for CAA* (definite/probable) | 3/43 |

Abbreviations: CAA, cerebral amyloid angiopathy. CAA-he, cerebral amyloid angiopathy related intracerebral hemorrhage. CAA-ri, cerebral amyloid angiopathy-related inflammation. ICH, intracerebral hemorrhage. cSS, cortical superficial siderosis. <sup>a</sup>all with clinical and radiological improvement. <sup>b</sup>one patient was lost during follow-up

**Table II.**
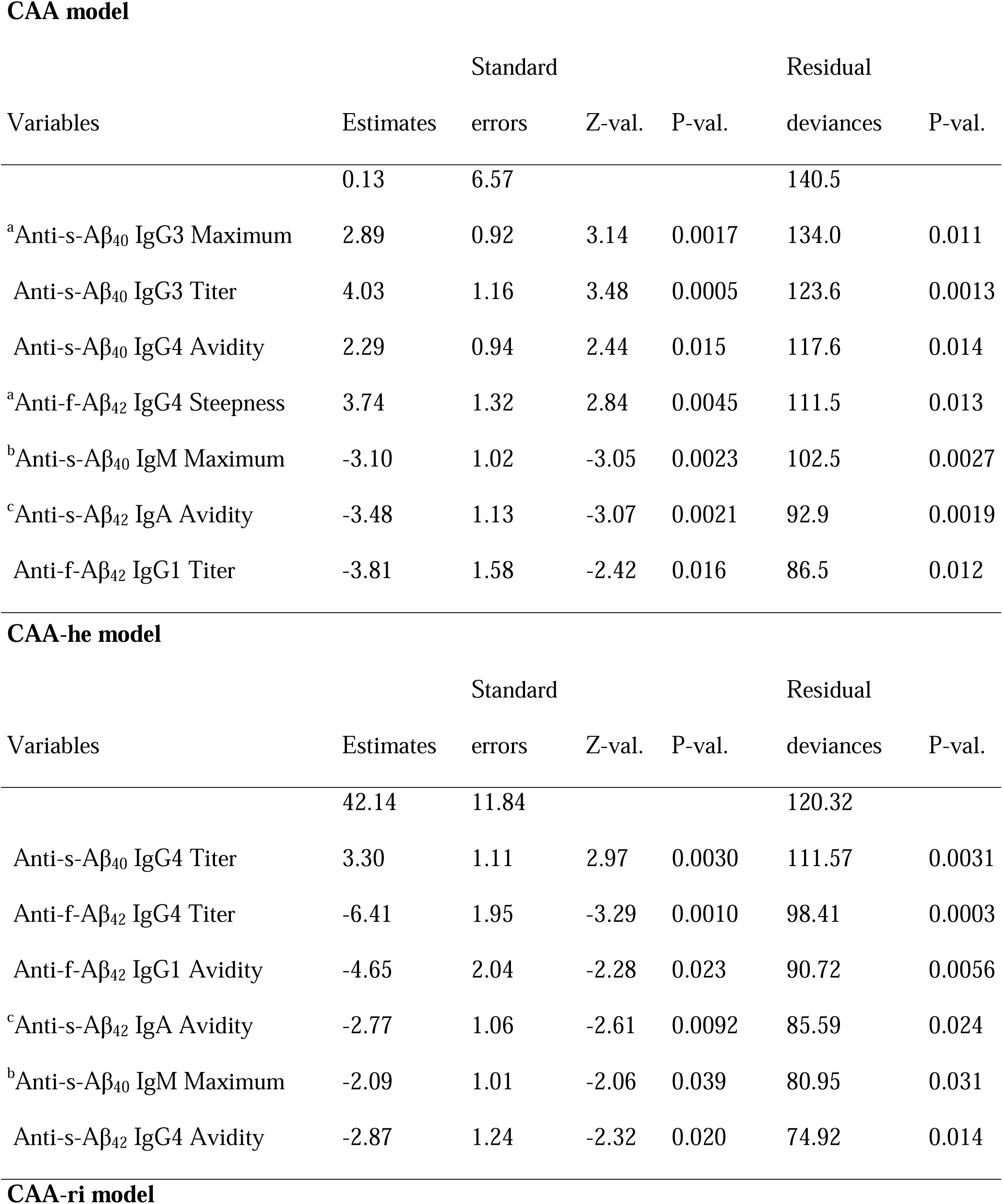
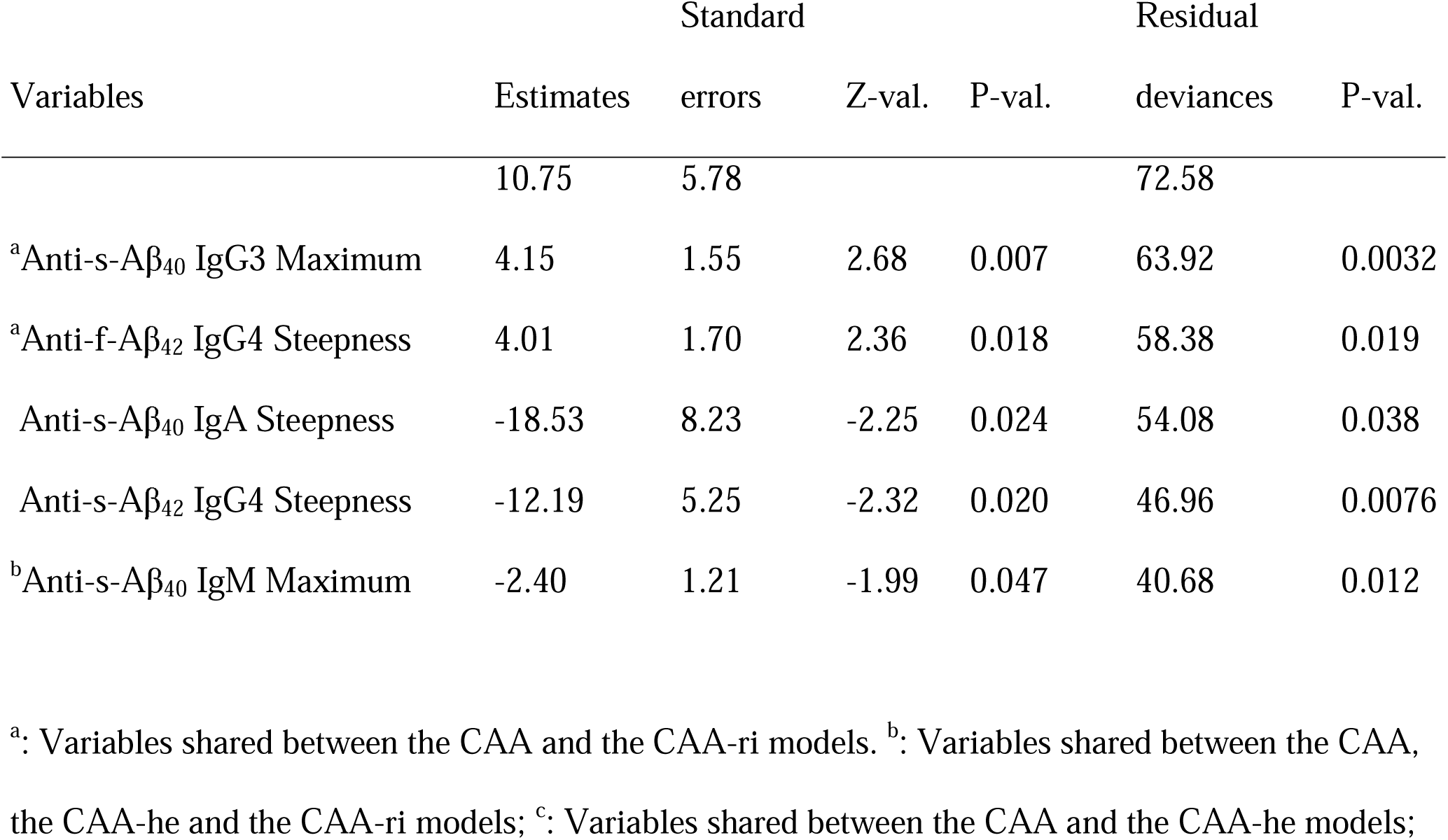
Multivariable logistic regression models for CAA, CAA-he, and CAA-ri against healthy aged controls CAA model

**Figure 2.**
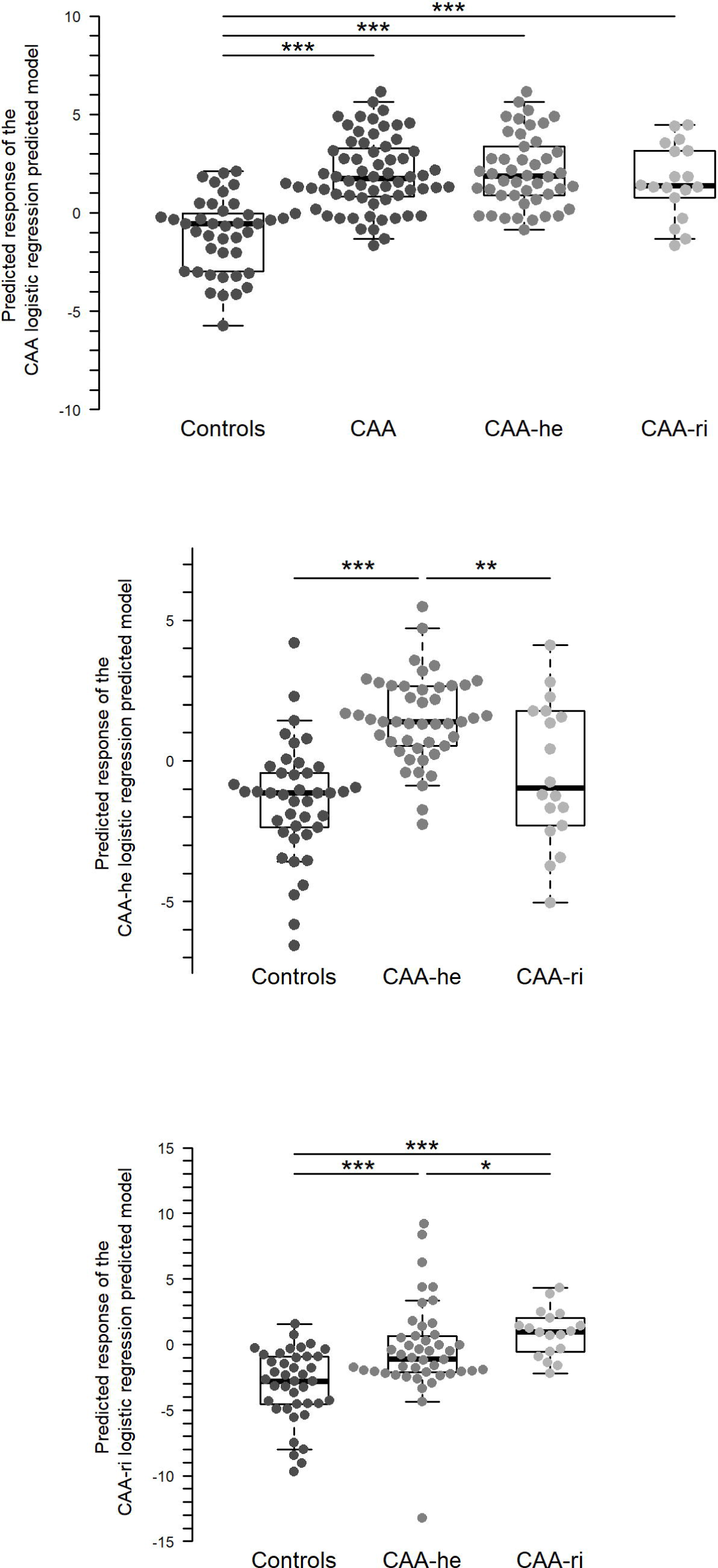
Serological differences associated with CAA clinical phenotypes. A, CAA-model predicted value using the logistic multivariable regression model presented in Table 2, upper part. B, CAA-he model predicted values using the logistic multivariable regression model presented in Table 2, middle part. C, CAA-ri model predicted values using the logistic multivariable regression model presented in Table 2, lower part. *:p<0.05; **: p<0.01; ***: p<0.001. Wilcoxon’s test.

The serological anti-Aβ antibody profile associated with CAA-he as compared to healthy aged controls (Table 2 and Fig 2B) also included lower diversity of anti-s-Aβ_40_ IgM and lower avidity of anti-s-Aβ_42_ IgA. Regarding the IgG4 isotype, CAA-he patients displayed higher concentrations of anti-s-Aβ_40_ IgG4, and lower concentrations and avidity of anti-Aβ_42_ IgG4, respectively. CAA-he patients also displayed lower avidity of anti-f-Aβ_42_ IgG1 as compared with healthy aged controls. Of note, excluding the 8 CAA-he patients that presented cSS without ICH did not change variables present in this model. All these variables and independently contributed to the model, without multicollinearity (all VIFs < 1.3).

As found in the CAA model, CAA-ri patients (Table 2 and Fig 2C) displayed lower diversity of anti-s-Aβ_40_ IgM, as also found for CAA-he patients, but also a higher diversity of anti-s-Aβ_40_ IgG3, and a higher steepness of anti-fibrillar Aβ_42_ IgG4 dilution curve. Conversely, CAA-ri patients displayed a lower steepness of anti-s-Aβ_42_ IgG4 and anti-s-Aβ_40_ IgA. All these variables contributed independently to the model, without multicollinearity (all VIFs < 1.4).

## Discussion

Analyses of blood anti-Aβ antibodies demonstrate complex serological profiles in CAA, displaying distinctive features in CAA-he and CAA-ri patients. This proof-of-concept study suggests: i) evidence of a link between anti-Aβ antibody responses and CAA; ii) existence of defined circulating anti-Aβ antibody species associated with distinct pathological phenotypes. In brief, clinical manifestations of CAA appear to relate, at least in part, to a biased natural antibody repertoire or abnormal responses to pathological Aβ peptide. However, at this stage, these serological profiles may not be used as biomarkers.

The causal relevance of these observations remains to be elucidated. Different naturally occurring anti-Aβ repertoires could be susceptibility factors for developing CAA, possibly by interfering with the clearance of cerebral Aβ. Cerebrovascular Aβ deposits may also induce anti-Aβ auto-immune responses emerging from the natural antibody repertoire. Finally, such induced anti-Aβ antibody species might enhance CAA and/or trigger hemorrhagic or inflammatory manifestations, as suggested in experimental mouse models.^7,8^

Although this multivariable modeling approach does not allow pathophysiological conclusions about the role of given anti-Aβ species in CAA, it suggests hypotheses and drives attention towards potentially relevant anti-Aβ features. With the notable exception of anti-fibrillar Aβ_1-42_ IgG4, all serologic parameters relating to Aβ_1-42_ were lower in CAA. Regarding antibodies reacting with Aβ_1-40_, our results show preferential developments of IgG3 and IgG4 antibody responses. Interestingly, lower diversity of anti-soluble Aβ_1-40_ IgM was the only common feature of CAA-he and CAA-ri patients. This could indicate an IgM response preferentially directed toward some particular pathogenic Aβ_1-40_ epitopes in response to Aβ_1-40_ cerebrovascular deposits.

The role of antibody isotypes in anti-Aβ antibodies related CAA manifestations is elusive. In AD patients treated with monoclonal anti-Aβ antibodies, amyloid related imaging abnormalities with vasogenic edema (ARIA-E) or with hemorrhagic features (ARIA-H) were initially reported with antibodies of IgG1 subclass.^22-24^ No ARIA-E but ARIA-H were yet reported in patients receiving anti-Aβ IgG4 (Crenezumab),^25^ while the existence of both remains uncertain for IgG2 (Ponezumab)^26,27^ None of the monoclonal anti-Aβ antibodies used in AD immunotherapy trials was of the IgG3 subclass. Like IgG1, IgG3 antibodies display potent effector functions including complement classical pathway activation and phagocytic and cytotoxic cell activation. IgG3 were thought to be more pathogenic than other IgG subclasses in anti-neutrophil cytoplasm antibody (ANCA)-associated vasculitis.^28^

Lobar ICH, MH and cSS were also reported in active immunotherapy trial CAD106, designed for eliciting Aβ-specific antibodies without T-cell response. Lobar ICH occurred in a patient without anti-Aβ IgG response to CAD106, but his anti-Aβ IgM status was not reported.^29^ It is worth noting that in ANCA-associated vasculitis, the presence of transient but recurrent IgM-ANCA is associated with a higher severity mainly due to acute hemorrhagic pulmonary manifestations.^30^

The main risks factors for CAA are age, coincidental Alzheimer’s disease, and ApoE genotypes, with controversy regarding the respective roles of ε2 and ε4 alleles.^31^ We accounted for age and cognitive decline by selecting aged healthy controls without cognitive decline, and CAA patients with normal MMSE. One potential limitation is that, due to unavailable material, ApoE genotypes could not be evaluated in this study. However, used diagnostic criteria for CAA-he and CAA-ri are independent of the ApoE genotype. Part of the serologic diversity might relate to ApoE genotypes, since both ε2^32,33^ and ε4^34^ alleles have been linked to more severe CAA, with vasculopathies and hemorrhagic phenotypes. This question should be addressed in further studies.

In previous studies, serum anti-Aβ antibodies were mostly analyzed in the context of Alzheimer’s disease, with various results reported by different authors using distinct methods.^12-14^ Neuropathological CAA is virtually present in all patients with AD pathology.^35^ However, lobar microbleeds evoking CAA are present in only about 20% AD patients^36^ while lobar ICH and spontaneous manifestations similar to CAA-ri are rare in AD. This may be explained by the distinct CAA neuropathological phenotypes in AD and in patients with CAA clinical manifestations.^37^ Whether AD patients with and without cerebral microbleed would present with different anti-Aβ serological signatures as compared to CAA-he and CAA-ri patients is thus likely and will be addressed in further studies.

Anti-Aβ antibodies circulate mostly as immune complexes,^11^ but whether these complexes include circulating Aβ, cross-reactive proteins, and/or anti-idiotype antibodies is not known. As previously performed by others,^11-12,38^ we used acidic dissociation of serum samples followed by extemporaneous neutralization to analyze all circulating anti-Aβ antibodies regardless of their free or bound state. We used high purity synthetic Aβ preparations and appropriate blank controls to ensure binding specificity. Of note, given the low specific signal obtained with non-dissociated samples once the background is subtracted, dilution curve analysis could not have been performed on non-dissociated samples. That anti-Aβ antibodies circulate mostly as immune complexes also raises questions regarding plasma Aβ levels measurements. Contradictory results exist on circulating Aβ levels in CAA.^39,40^ The circulating levels of free Aβ should depend on the concentrations and affinities of circulating anti-Aβ antibodies, and it is possible that the experimental conditions in which Aβ measurements are performed cannot fully circumvent Aβ masking by circulating anti-Aβ antibodies.

In conclusion, this correlative proof-of-concept study demonstrates distinct serum anti-Aβ antibody patterns in CAA and its hemorrhagic and inflammatory manifestations. Larger prospective and experimental studies could elucidate the triggering role of anti-Aβ antibodies in spontaneous or immunotherapy-induced CAA manifestations, and provide appropriate biomarkers.

## Data Availability

Data are available upon reasonable request.

## Acknowledgments

We thank Dr Margaret Goodall (Birmigham, UK) for providing anti-IgG subclass antibodies.

## Author contributions

Study concept and design: YC, JC, SA, PA. Data acquisition and analysis: YC, JC, DD, JF, MF, FK, TC, TA, XA, ZB, TdB, CC, GT, RP, AW, MS, DR, CC. Drafting the manuscript and figures: YC, JC, GD, CC, SA, PA.

